# Health Complexity Assessment in Primary Care: a validity and feasibility study of the INTERMED tool

**DOI:** 10.1101/2020.10.21.20216929

**Authors:** Camila Almeida de Oliveira, Bernardete Weber, Jair Lício Ferreira dos Santos, Miriane Lucindo Zucoloto, Lisa Laredo de Camargo, Ana Carolina Guidorizzi Zanetti, Magdalena Rzewuska, João Mazzoncini de Azevedo-Marques

## Abstract

**Background:** While considerable attention has been devoted to patients’ health complexity epidemiology, comparatively less attention has been paid to tools to identify and describe, in a personalized and comprehensive way, “complex patients” in primary health care (PHC).

**Objective:** To evaluate INTERMED tool’s validity and feasibility to assess health complexity in PHC.

**Design:** Cross-sectional psychometric study.

**Setting:** Three Brazilian PHC Units.

**Participants:** 230 patients above 18 years of both sexes.

**Measurements:** Spearman’s rho assessed concurrent validity between the whole INTERMED and their four domains (biological, psychological, social, health system) with other well-validated instruments. Pearson’s X^2^ measured associations of the sum of INTERMED “current state” items with use of PHC, other health services and medications. Cronbach’s Alpha assessed internal consistency. INTERMED acceptability was measured through patients’ views on questions and answers’ understanding and application length as well as objective application length. Applicability was measured through patients’ views on its relevance to describe health aspects essential to care and INTERMED’s items-related information already existing in patients’ health records.

**Results:** 18.3% of the patients were “complex” (INTERMED’s 20/21 cut-off). Spearman’s correlations located between 0.44 - 0.65. Pearson’s coefficients found were X^2^ = 26.812 and X^2^ = 26.883 (both p = 0.020) and X2 = 28.270 (p = 0.013). Cronbach’s Alpha was 0.802. All patients’ views were very favorable. Median application time was 7 minutes and 90% of the INTERMED’s interviews took up to 14 minutes. Only the biological domain had all its items described in more than 50% of the health records.

**Limitations:** We utilized the cutoff point used in all previous studies, found in research performed in specialized health services.

**Conclusion:** We found good feasibility (acceptability and applicability), and validity measures comparable to those found from specialized health services. Further investigations of INTERMED predictive validity and suitability for routine PHC use are worthwhile.

## Introduction

To deliver better healthcare for each person, Primary Health Care (PHC) teams have progressively broadened their structure and work processes in response to the need to handle entangled biological, social and psychological aspects of health (1–3). An assessment of that biopsychosocial complexity (4,5) can identify aspects of health related to a person (e.g. chronic disorders and symptoms that interfere with functionality; emotional state, lifestyle, and social context) and their health services (e.g. organization and coordination, service access, treatment experience) (4,6,7). Such knowledge can help to target those in need for advanced care planning, and effectively develop, coordinate and monitor those plans (1,7,8). Considerable attention has been devoted to identifying the epidemiology of patient complexity, predominantly focusing on medical diagnoses (number of chronic disease and severities) (9). However, comparatively less attention has been paid to how to effect the identification of ‘complex patients’, incorporating not only the physical condition but the mental health and social needs (10), who need care planning in the clinical routine of PHC services (1,4,5).

INTERMED Complexity Assessment Grid is a standardized instrument which objectives are to operationalize the assessment of the biopsychosocial complexity of a person’s health needs and improve the communication flow between professionals, patients and services (11–13). Its development was methodologically robust (14) as well as its validation in different patient populations, including in- and out-patients in secondary, tertiary and emergency services, with a range of health problems (11,12,15–18) and using different versions (face-to-face interview (19); self-assessment (20); pediatric (21), adults (19), and elderly (18)).

Longitudinal epidemiological studies showed that the INTERMED’s scores at baseline correlates with clinically meaningful indicators at follow-up, regarding hospital (22–24) and community (25–27) care as well as mortality (28), healthcare costs (27,29,30) and quality-of-life (29). INTERMED versions helped to construct person-centered general elderly profiles in two countries (31,32). Before-and-after studies (33) and randomized controlled trials (RCTs) (34–37) deployed INTERMED as a method of integrative health risk assessment in its design. Case managers utilize INTERMED versions to identify people with a specific profile of health needs complexity and coordinate integrated care for them (7,38). A recent systematic review that compared screening tools to identify patients with complex health needs at risk of high use of healthcare services, concluded that the INTERMED’s adult version is the most promising tool and hence recommended its use (6). However, to date, only one study assessed the psychometric properties of the INTERMED versions in PHC patients, involving a small sample (n=55) and only evaluating the concordance between the self-assessment version with the adult interview version as gold standard (39).

In this study, we aimed to evaluate the psychometric values of INTERMED Complexity Assessment Grid in a PHC adult population. We hypothesized that INTERMED could have adequate validity and feasibility (applicability and acceptability) in a PHC population, a fundamental condition for its use to categorize complexities that inform subsequent personalized comprehensive care plans in that setting.

## Methods

### Sampling and recruitment procedures

The sampling was based on the two types of Brazilian public organizational models of PHC, the “Basic Health Unit” (BHU) and the “Family Health Strategy” (FHS) model (40). Three PHC units located in the city of Ribeirão Preto (state of São Paulo, Southeast region of Brazil) participated in the study: one FHS service managed by Ribeirão Preto Medical School of University of São Paulo; and one FHS and one BHU managed by Municipal Health Secretary.

The sample was divided into 10 groups stratified by age (18-30, 31-40, 41-50, 51-60 and ≥60 years) and sex, with a minimum of 15 and maximum of 35 people, to obtain the representativeness of these different age and sex groups. Inclusion criteria were as follow: living in the catchment area of three PHC units, having health records, speaking, and understanding Portuguese, and attending a PHC unit at a moment of recruitment.

Participant’s recruitment took place when they were in a PHC unit in a queue awaiting attendance (for their own health). When a patient refused to participate in the study, a researcher invited the next consecutive patient. Voluntary written informed consent was obtained from each participant, during which a permission to review health records was obtained.

### Instruments

INTERMED Complexity Assessment Grid is a semi-structured interview, with 17 questions (14). It has four domains-biological, psychological, social, and health system - with 20 items, constituting a matrix formed by three temporal axes. Eight items are about health care before the current treatment episode (<u>“historical”</u>, with two items for each domain), eight items about current treatment episode (“<u>current state</u>”, with two items for each domain) and four items about prognoses of health care needs <u>(“vulnerability/prognoses”</u>, with one item for each domain). Each item in the different domains scored according to Clinical Anchor Points, ranging from zero (“no vulnerability or need”) to a score of three (“high vulnerability or need”) (11,12,14,19). The items can be summed up to a total score ranging from 0 to 60 and reflecting the level of complexity of the case (cut-off point of 20/21, derived from studies carried out in specialized services) (23). We used the Portuguese Brazilian version previously translated and validated with inpatients (41,42). The INTERMED was used both to assess the profile of the patients’ level of complexity through the interview and to analyze the data found in the health records (43).

Due to the pioneering character of INTERMED - that is, it was developed to assess health complexity in a broad, biopsychosocial way, which was not done in a structured way by any other instrument until then - there is no other instrument that can be considered as a standard-gold against which it can be valued (11,12,14). Therefore, as done in the other studies on INTERMED validation carried out previously, we evaluated the concurrent validity of each of its four domains with other already well-validated specific instruments for these domains, which are described below (11,12,14).

The other instruments used in this study were: 1) Socio-demographic Questionnaire developed for the study; 2) Hospital Anxiety and Depression Scale (HADS), with 14 items, each item is ranging from zero to three points (44); 3) Medical Outcomes Study – Social Support Survey (MOS-SSS), with 19 items ranging five points (45); 4) Charlson Comorbidity Index (CCI) to categorizing physical comorbidities, consisting of 19 selected conditions that are weighted from 1 to 6 and summed to an index on a 0–33 scale (46); 5) WHO Quality of Life – Bref (WHOQOL-BREF), with 26 questions separated into four domains (physical health, psychological health, social relationships, and the environment) (47), 6) Questionnaire for Evaluation of Health Services Use, developed for the purpose of this study through adaptation of the former Questionnaire from the SABE Study (48), focusing the patients’ use of PHC, other health services (hospitalization, emergency, and specialists) and medications in the last six months; and 7) Feasibility Questionnaire (49) for patients, developed for this study and with Likert scale questions (five response options each) focusing on: acceptability (satisfaction level from understanding the questions; satisfaction level from understanding the answers; satisfaction with the length of the interview); and applicability (the relevance of asking the questions within each of the four domains - biological, psychological, social and health system - to help with health care delivery). These seven items of the Feasibility Questionnaire had its five response possibilities grouped into satisfactory and unsatisfactory.

### Training researchers to apply INTERMED

The authors of the Brazilian Portuguese version of INTERMED (41,42) trained, during a three-day workshop, two researchers that collected the data (an occupational therapist (CAO) and a nurse (LLC)).

### Data collection procedures

The researchers collected the data from November 2018 to June 2019. To understand the operation and determine the order of data collection in the three health units, the researchers followed the routine of each service for a week; after this, the data collection took place in each health unit for two months, covering every day and period of operation (7 a.m to 5 p.m, Monday through Friday). The principal researcher (CAO) interviewed the patients using the listed instruments and the INTERMED had its interview length timed.

Study data were collected and managed using REDCap (Research Electronic Data Capture) (50), a web-based software platform data capture tool, hosted at the Ribeirão Preto Medical School of University of São Paulo (Department of Social Medicine) - https://research.fmrp.usp.br/.

### Health records review

Following semi-structured interviews, participants had their health records (both paper and electronic) reviewed by academic-clinicians (CAO, LLC)

### Statistical Analysis

To measure construct validity, we used Spearman’s rho to correlate the total score of the INTERMED and the scores of its three domains (biological, psychological, and social) with the scores of CCI, HADS and MOS-SSS respectively; and also with the total score of WHOQOL-brief, the scores of three of its domains (physical, psychological, social) and its question 24 (“How satisfied are you with your access to health services?”, to evaluate correlations with INTERMED’s health system domain). Coefficients ranging from 0.10 to < 0.40, from 0.40 to < 0.70, from 0.70 to < 1.00, were interpreted as weak, moderate an strong respectively (51).

We also used Pearson’s X^2^ (51) to compare the sum of the INTERMED’s eight “current state” variables of the four domains and the sum of the two “current state” variables of each domain with three variables of health service use (PHC, other health services, and medications’ use). Cronbach’s Alpha assessed internal consistency of INTERMED and values ranging from < 0.5, from 0.5 to 0.6, from 0.6 to 0.7, from 0.7 to 0.8, from 0.8 to 0.9 and from ≥ 0.9, were interpreted as unacceptable, poor, questionable, acceptable, good and excellent (52).

The feasibility analysis involved estimation of: distribution of absolute and relative frequencies of “satisfactory” answers for each of the seven topics asked patients in the Feasibility Questionnaire (acceptability and applicability); the objective INTERMED application time in the interview with the patient (acceptability); and the presence of information on each of the INTERMED variables in the medical records (43) (applicability). Analysis was conducted using IBM SPSS Statistics for Windows, version 20.0.

### Ethics approval

The Research Ethics Committee of the Community Health Center of the Ribeirão Preto Medical School of the University of São Paulo approved the study (n° 99566718.0.0000.5414 in 10/2018).

## Results

Two hundred and thirty-eight (238) patients agreed to participate; eight did not complete the interview due to lack of time. Table 1 shows socio-demographic characteristics of the 230 patients (mean age = 45.92 (±15.43) years, 56.1% female, 53.5% reported being white).

**Table 1.**
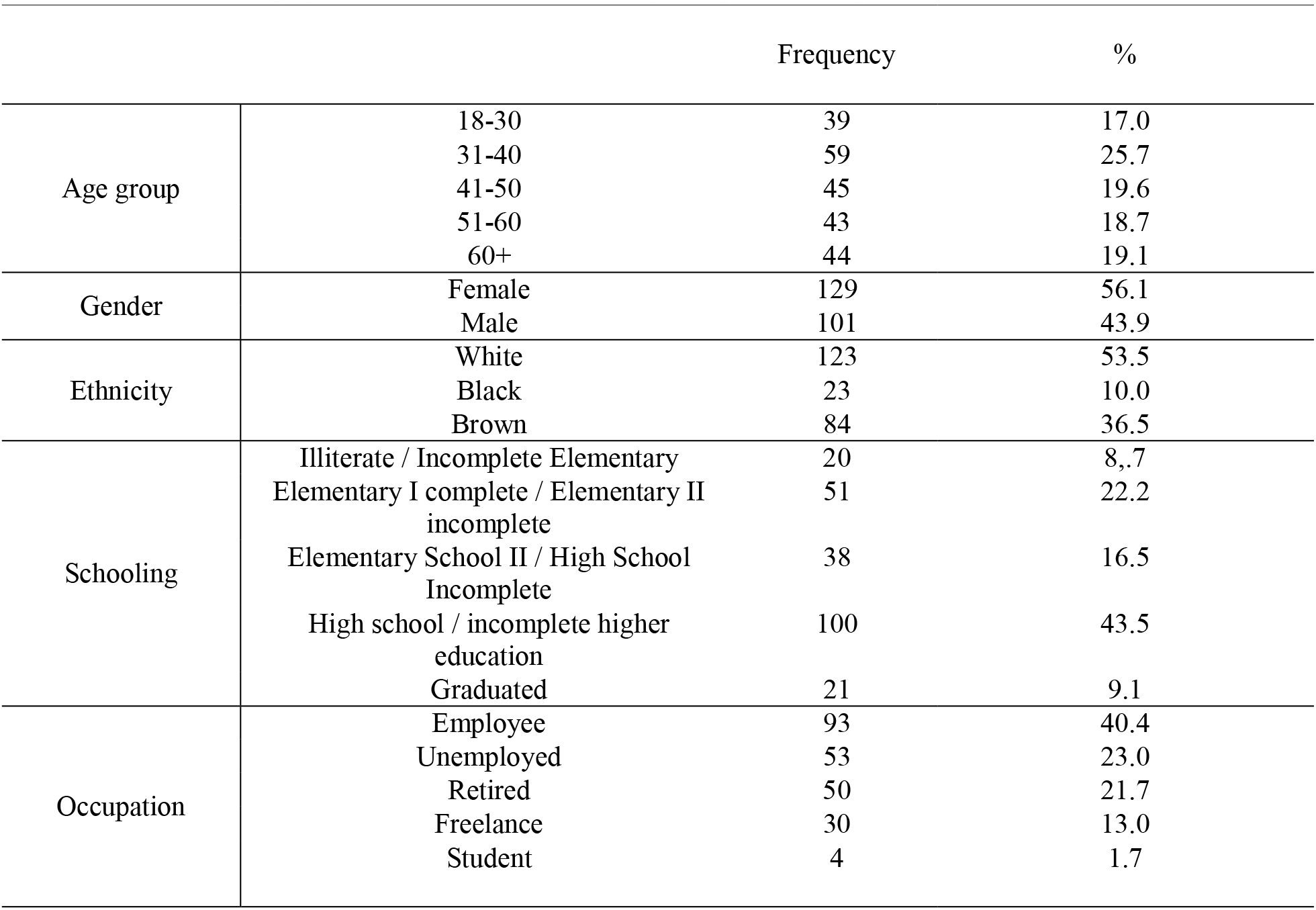
Socio-demographic characteristics of the 230 participants, PHC patients.

Table 2 describes INTERMED profiles of the participants according Clinical Anchor Points of each variable. INTERMED minimum total score value was zero, maximum value was 38, mean was 13.57 and median was 13. Forty-two (18.3%) in the total sample were “complex”, according to the 20/21 cut-off score (23). Ninety-two patients (40.0% of the total sample) presented physical-mental multimorbidity (score 2 or 3 in the variable “Chronicity” of INTERMED and score 2 or 3 in the category “Psychiatric symptoms”); of these, 34 (14.8%) were classified as complex. Thirty-two (13.9%) had only physical multimorbidity (only a score of 3 in “Chronicity”), of which 2 (0.9%) were complex.

**Table 2.**
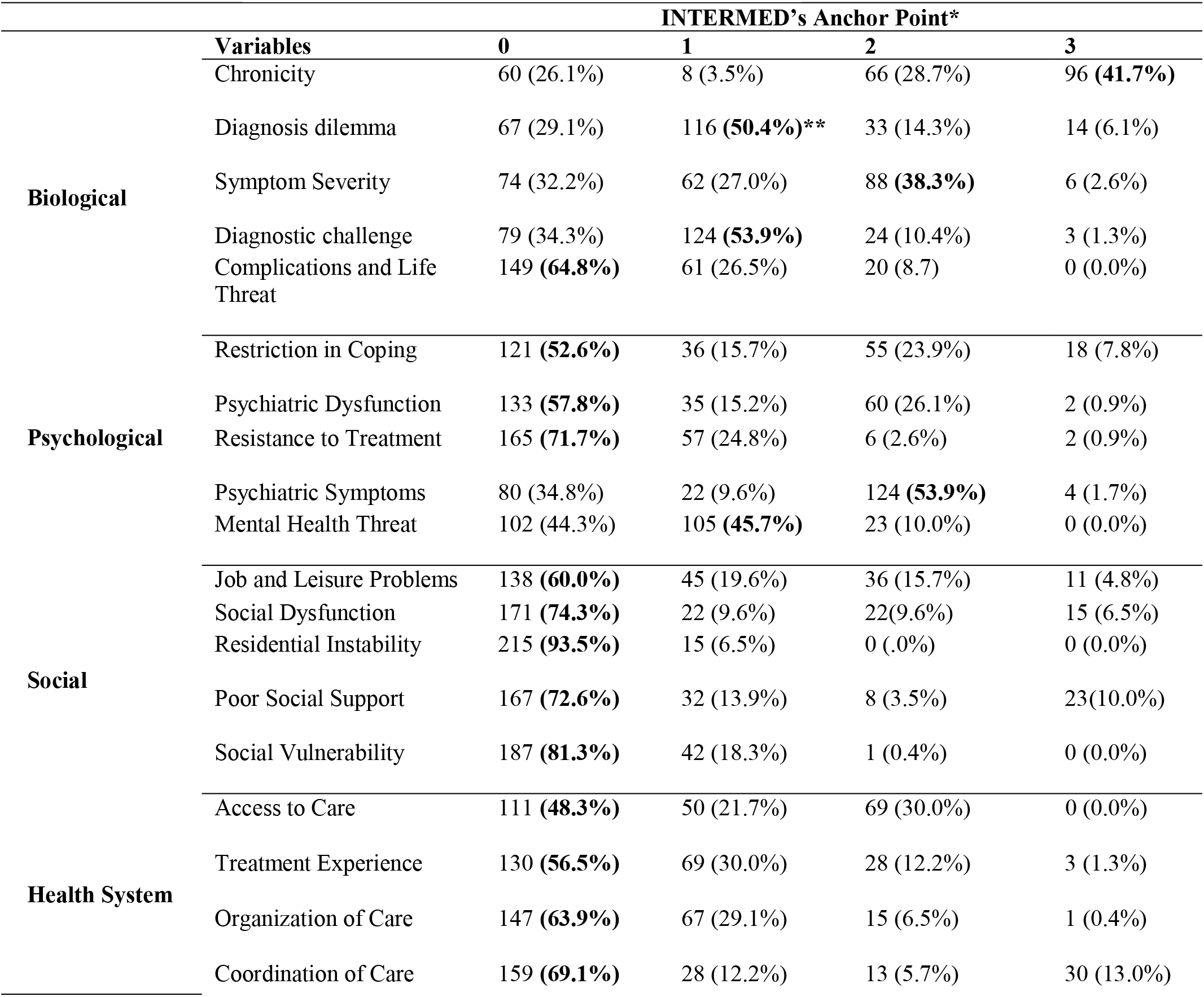
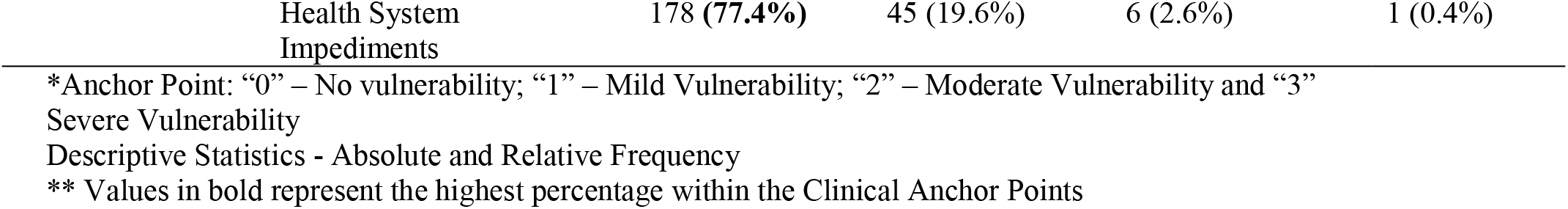
Profiles of the 230 PHC patients regarding INTERMED items and their Anchor Points.

### Validity

Regarding concurrent validity, Spearman’s correlation coefficients, between INTERMED and HADS, MOS-SS, CCI, and WHOQOL-BREF scores, ranged from 0.44 to 0.65, suggesting moderate concordance (51) (Table 3).

**Table 3.** Spearman’s correlation coefficients between INTERMED and other tools.

|  | Biological | Psychological | Social | Health System | INTERMED total |
| --- | --- | --- | --- | --- | --- |
| HADS: |  |  |  |  |  |
| Total | 0.30 | **0.59** | 0.40 | 0.26 | 0.55 |
| Anxiety | 0.33 | **0.57** | 0.37 | 0.22 | 0.54 |
| Depression | 0.20 | **0.50** | 0.36 | 0.26 | 0.46 |
| MOS - SSS | -0.16 | -0.35 | **-0.44** | -0.17 | -0.38 |
| CCI | **0.47** | 0.13 | 0.11 | 0.08 | 0.30 |
| WHOQOL-BREF: |  |  |  |  |  |
| Total | -0.37 | -0.52 | -0.36 | -0.37 | **-0.57** |
| Biological | -0.61 | -0.53 | -0.33 | -0.29 | **-0.65** |
| Psychological | -0.25 | **-0.49** | -0.36 | -0.19 | -0.44 |
| Social | -0.24 | -0.41 | -0.37 | -0.21 | **-0.42** |
| *Question 24 | **-0.43** | -0.15 | -0.06 | -0.38 | -0.20 |

HADS = Hospital Anxiety and Depression Scale; MOS-SSS = Medical Outcomes Study – Social Support Survey; CCI = Charlson Comorbidity Index; WHOQOL-BREF = World Health Organization Quality of Life – Bref.
\*Question 24 - "How satisfied are you with your access to health services?"

Cronbach’s Alpha was 0.802, a good internal consistency’s value (52). After deleting each of the 20 items from INTERMED, the Cronbach’s Alpha values found were located between 0.781 and 0.810. Three items showed no decrease in the original Cronbach’s Alpha value when deleted: “Treatment Experience” and “Resistance to Treatment” (deletion of each one increased Cronbach’s Alpha to 0.806); and “Job and Leisure Problems” (0.810).

Pearson coefficients (X^2^) for health services use and the sum of INTERMED’s “current state” items are presented in Table 4. The sum of the scores of all INTERMED’s” current state” items showed a significant association with both the use, in the last six months, of PHC, other health services, and medications (p < 0.005).

**Table 4.** Association between the Current State INTERMED context and the use of the health service.

| Health Service Use* | Temporal Context | Current State Domain |  |  |  |
| --- | --- | --- | --- | --- | --- |
|  | Current State total*** | Biological | Psychological | Social | Health System |
| **Other health services | $X^2 = 28.270$,<br>$p = 0.013$ | $X^2 = 10.759$,<br>$p = 0.056$ | $X^2 = 3.067$,<br>$p = 0.690$ | $X^2 = 5.120$,<br>$p = 0.275$ | $X^2 = 8.707$,<br>$p = 0.121$ |
| PHC | $X^2 = 26.812$,<br>$p = 0.020$ | $X^2 = 17.777$,<br>$p = 0.003$ | $X^2 = 2.635$,<br>$p = 0.756$ | $X^2 = 4.930$,<br>$p = 0.295$ | $X^2 = 6.176$,<br>$p = 0.289$ |
| Use of medication | $X^2 = 26.883$,<br>$p = 0.020$ | $X^2 = 21.521$,<br>$p = 0.001$ | $X^2 = 9.125$,<br>$p = 0.104$ | $X^2 = 15.286$,<br>$p = 0.004$ | $X^2 = 12.860$,<br>$p = 0.025$ |

\*Health Service Use - the absolute number of appointments that the patient used, and the amount of medication used
\*\* Others health services – number of hospitalizations, visits of emergency room, appointments with specialists and clinics (physical and mental)
\*\*\* Current State total - sum of the results of the eight variables of the current state context
Descriptive Statistics – Pearson $X^2$ , p-value threshold ( $P < 0.005$ ) for a statistical significance

### Feasibility

Regarding acceptability, all participants understood the questions and answers and were satisfied with the application length; and concerning objective application length, mean was 8.51 minutes, median 7 minutes, and up 14 and 18.15 minutes for 90% and 95% of the patients, respectively. Regarding the applicability, concerning the relevance of information from INTERMED’s domains to help in healthcare, the numbers of patients who found them relevant were very high across all domains (biological (n=230, 100%); psychological (n=227, 98.7%); health system (n=221, 96.1%); and social (n= 215, 93.5%)). As for applicability, the health records review (Table 5) showed that the INTERMED’s domains “social” and “health system” were the least likely to have complete data in the health records, particularly items “Experience of treatment” and “Social support”. In contrast, the biological domain had the most complete information across all its variables. It was not possible to calculate the total score of INTERMED from health records for any participant, due to a lack of required information. These data showed that INTERMED had clinical relevance regarding obtaining (and presenting in a practical way) data that were not available through the health records already existing in PHC.

**Table 5.** Completeness of INTERMED’s Domains in the Health Records.

| Domain | INTERMED variables* | Availability in health records<br>(Frequency, %) |
| --- | --- | --- |
| Biological | Chronicity | 211 (88.7%) |
|  | Diagnosis dilemma | 202 (84.9%) |
|  | Diagnostic challenge | 194 (81.5%) |
|  | Complications and Life Threat | 136 (57.1%) |
|  | Symptom Severity | 122 (51.3%) |
| Psychological | Resistance to Treatment | 153 (64.3%) |
|  | Restriction in Coping | 84 (35.3%) |
|  | Psychiatric Symptoms | 81 (34.0%) |
|  | Mental Health Threat | 80 (33.6%) |
|  | Psychiatric Dysfunction | 39 (16.4%) |
| Social | Job and Leisure Problems | 48 (20.2%) |
|  | Social Dysfunction | 42 (17.6%) |
|  | Residential Instability | 19 (8.0%) |
|  | Social Vulnerability | 16 (6.7%) |
|  | Poor Social Support | 10 (4.2%) |
| Health System | Organization of Care | 180 (75.6%) |
|  | Coordination of Care | 78 (32.8%) |
|  | Health System Impediments | 51 (21.4%) |
|  | Access to Care | 25 (10.5%) |
|  | Treatment Experience | 3 (1.3%) |

\* Order of frequency of availability
Descriptive Statistics I - Absolute and Relative Frequency

## Discussion

We explored validity and feasibility of the INTERMED adult interview tool applied in PHC’s attendees in Brazil, using an adequate sample size, multiple performance metrics, and patients’ opinions. To our knowledge, this is the second in general, and a first such a comprehensive evaluation of INTERMED in a PHC population.

We found moderate Spearman’s correlations (51) with other instruments related to the four INTERMED domains, as in research from tertiary services (ranging from 0.55 to 0.74) (15). This can be linked to the fact that the goal of INTERMED is to enable a wider detection and description of individual health care needs; and not to collect specific diagnostic data. The relationship of these health care needs to specific diagnostic data can be detailed as its results are used to assist in construction and monitoring of personalized health care plans (14).

A good Cronbach’s alpha value (52) of 0.802 was found, akin to those from tertiary populations (ranging from 0.8 and 0.89) (15,53); meaning that INTERMED presents an adequate internal consistency also in that PHC setting (49,52). The significant association between INTERMED’s “current status” scores and patients’ use of PHC, other health services and medications demonstrates that it correlates with clinically significant variables for care planning and monitoring in PHC (9,10,54). Previous studies, assessing patients’ health complexity by INTERMED, have demonstrated a positive association between its scores and increased risk of hospitalization and health care costs (19,26).

Regarding feasibility, patients felt the tool captured relevant information about their health care needs, which, otherwise were rarely described in their health records, except for those related to the biological domain. This last finding is a widely reported problem and a target for improvement efforts (55,56). Previous research suggests that health records with systematized comprehensive (biopsychosocial) data allows greater integration of care through better communication between patients, professionals and health services (57,58); and facilitate delivery of evidence-informed personalized health care plans (59). Our findings does not mean that health care providers (HCPs) of the studied sample do not attend to the complex psychosocial circumstances of their patients, but rather that INTERMED could help to prompt PHC teams to assess all relevant aspects and make the process of capturing and sharing these data more effectively to assist in the development of personalized health care plans, informed by the individual profiles generated by the Clinical Anchor Points of each INTERMED variable (19). The maximum 14 minutes needed to complete the interview for 90% of the patients, is shorter than the recommended duration of a single outpatient appointment in Brazil (60).. The median of the length time – 7 minutes – is compatible with the average appointment duration practiced by PHC physicians in 39 countries (61). Together with all patients feeling satisfied with the application time, these findings suggest that INTERMED, as a practical personalized assessment tool, presents an opportunity to overcome the challenge of assessing complexity routinely during PHC appointments (3,62– 69).

This study has a few important implications. With nearly half of the sample being multimorbid, yet overall 18.3% found to be “complex”, when adopting a personalized-biopsychosocial lens, this study supports the notion that in practical terms defining PHC patient complexity as morbidity alone is inadequate(4–6,69). Instead, we support an increasingly recommended PHC person-centered planning including biopsychosocial (comprehensive) health risk assessment(1,8,55,56,62–68), which can facilitate an better integration of care and a decrease in the cost of the health network(10,70). INTERMED could enable this approach once its predictive validity and feasibility from PHC teams’ perspective is confirmed. In fact, RCTs and implementation efforts to successfully deliver such a care planning in PHC have recently been conducted(71–73) or are under development(74), including one RCT that used the INTERMED tool (35)

Further investigations are also needed to establish how, integrated on centralized electronic health records (EHS) (75) and telemedicine (76,77), INTERMED could become a component of a computerized health decision support tool, profile-guided and action-oriented, to deliver personalized care planning in health services networks (78,79). In this way, INTERMED could to help direct health system networks towards the much-needed biopsychosocial, people-centered and integrated care, including in Brazil and other low-and-middle income countries (80–84).

### Study Limitations

While we utilized the cutoff point used in all previous studies, it would be important to research cutoff points with specific clinical significance for PHC. Correlating the categorization based on the INTERMED score with the clinical assessment of the PHC teams and applying clustering methods capable of handling many variables in larger PHC samples are two ways that are worth to verify.

## Conclusions

The data suggest that INTERMED has adequate psychometric properties to help PHC teams to assess the biopsychosocial complexity of health needs. INTERMED could assist PHC professionals and teams in defining patient group profiles and in the development of healthcare plans. The results indicate the need for further studies to assess the INTERMED’s potentialities to help the delivery of comprehensive, integrated, and person-centered care.

## Data Availability

As the data presents sensitive information that can facilitate the recognition of specific people, they are not in public repositories, but can be requested for the first case

## Abbreviations

BHU: Basic Health Unit
CAO: Camila Almeida de Oliveira
CCI: Charlson Comorbidity Index
FHS: Family Health Strategy
HADS: Hospital Anxiety and Depression Scale
HCP: Health Care Provider
LLC: Lisa Laredo de Camargo
MOS-SSS: Medical Outcomes Study – Social Support Survey
PHC: Primary Health Care
RCT: Randomized Controlled Trial
REDCap: Research Electronic Data Capture
WHOQOL-BREF: WHO Quality of Life – Bref

## Acknowledgments

We would like to thank Professor Craig Ramsay for his comments on this work during the international meeting held at the Health Services Research Unit of the University of Aberdeen in July 2019.

## Funding

This study was financed in part by the “Coordination of Superior Level Staff Improvement – Brazil (CAPES) – Finance Code 001”. It was also funded by the “Foundation for Support to Teaching, Research and Assistance at Clinics Hospital of Ribeirão Preto Medical School of University of São Paulo – Brazil (FAEPA)”. A discussion regarding it occurred during an international meeting funded by the “Global Challenge Research Fund (GCRF) - Internal Pump Priming Fund Round 5 of the University of Aberdeen”. The funding sources had no role in the design, conduct, and reporting of the study.

## Conflict of Interest Disclosures

The authors declare that they have no conflict of interest.

## Notes

### Competing Interest Statement

The authors have declared no competing interest.

### Author Declarations

The Research Ethics Committee of the Community Health Center of the Ribeirao Preto Medical School of the University of Sao Paulo approved the study (n 99566718.0.0000.5414 in 10/2018).

## References

1. Coulter A, Entwistle VA, Eccles A, Ryan S, Shepperd S, Perera R. Personalised care planning for adults with chronic or long-term health conditions. Cochrane Consumers and Communication Group, editor. Cochrane Database of Systematic Reviews [Internet]. 2015 Mar 3 [cited 2020 May 12]; Available from: http://doi.wiley.com/10.1002/14651858.CD010523.pub2

2. Swankoski KE, Peikes DN, Palakal M, Duda N, Day TJ. Primary Care Practice Transformation Introduces Different Staff Roles. The Annals of Family Medicine. 2020 May;18(3):227–34.

3. Baird B, Reeve H, Shilpa R, Honeyma M, Nosa-Ehima M, Sahib B, et al. Innovative models of general practice [Internet]. 2018 [cited 2020 Apr 16]. Available from: www.kingsfund.org.uk/publications/innovative-models-general-practice

4. Marcoux V, Chouinard M-C, Diadiou F, Dufour I, Hudon C. Screening tools to identify patients with complex health needs at risk of high use of health care services: A scoping review. Virgili G, editor. PLOS ONE. 2017 Nov 30;12(11):1–14.

5. Schaink AK, Kuluski K, Lyons RF, Fortin M, Jadad AR, Upshur R, et al. A Scoping Review and Thematic Classification of Patient Complexity: Offering a Unifying Framework. Journal of Comorbidity. 2012 Jan;2(1):1–9.

6. Lee L, Patel T, Hillier L, Locklin J, Milligan J, Pefanis J, et al. Frailty Screening and Case-Finding for Complex Chronic Conditions in Older Adults in Primary Care. Geriatrics. 2018 Jul 7;3(3):39.

7. Kathol RG, Hobbs Knutson K, Dehnel PJ. Physician’s Guide understanding and working with Integrated Case Managers [Internet]. Cham: Springer International Publishing: Imprint: Springer; 2016 [cited 2019 May 13]. Available from: http://dx.doi.org/10.1007/978-3-319-28959-5

8. Roberts S, Eaton S, Finch T, Lewis-Barned N, Lhussier M, Oliver L, et al. The Year of Care approach: developing a model and delivery programme for care and support planning in long term conditions within general practice. BMC Family Practice [Internet]. 2019 Dec [cited 2020 May 12];20(1). Available from: https://bmcfampract.biomedcentral.com/articles/10.1186/s12875-019-1042-4

9. Hwang AS, Atlas SJ, Hong J, Ashburner JM, Zai AH, Grant RW, et al. Defining Team Effort Involved in Patient Care from the Primary Care Physician’s Perspective. Journal of General Internal Medicine. 2017 Mar;32(3):269–76.

10. Bolen SD, Stange KC. Investing in Relationships and Teams to Support Managing Complexity. Journal of General Internal Medicine. 2017 Mar;32(3):241–2.

11. de Jonge P, Huyse FJ, Stiefel FC, Slaets JPJ, Gans ROB. INTERMED—A Clinical Instrument for Biopsychosocial Assessment. Psychosomatics. 2001 Mar;42(2):106–9.

12. Huyse FJ. From consultation to complexity of care prediction and health service needs assessment. Journal of Psychosomatic Research. 1997 Sep;43(3):233–40.

13. Intermed Foundation. INTERMED Complexity Assessment Grid (IM CAG version 6). Intermed Foundation; 2009.

14. Stiefel FC, Huyse FJ, Söllner W, Slaets JPJ, Lyons JS, Latour CHM, et al. Operationalizing Integrated Care on a Clinical Level: the INTERMED Project. Medical Clinics of North America. 2006 Jul;90(4):713–58.

15. de Jonge P, Hoogervorst ELJ, Huyse FJ, Polman CH. INTERMED: a measure of biopsychosocial case complexity: one year stability in Multiple Sclerosis patients. General Hospital Psychiatry. 2004 Mar;26(2):147–52.

16. Intermed Foundation. Manual for interpreting the INTERMED Self-Assessment(IMSA). Intermed Foundation; 2017.

17. Nader A. PEDIATRIC-INTERMED© COMPLEXITY ASSESSMENT GRID MANUAL for Inflammatory Bowel Disease (pIBD-INTERMED). Intermed Foundation; 2020.

18. Peters LL, Boter H, Slaets JPJ, Buskens E. Development and measurement properties of the self assessment version of the INTERMED for the elderly to assess case complexity. Journal of Psychosomatic Research. 2013 Jun;74(6):518–22.

19. Huyse FJ, Lyons JS, Stiefel FC, Slaets JP, de Jonge P, Fink P, et al. “INTERMED”: a method to assess health service needs. I. Development and reliability. Gen Hosp Psychiatry. 1999 Feb;21(1):39–48.

20. Boehlen FH, Joos A, Bergmann F, Stiefel F, Eichenlaub J, Ferrari S, et al. [Evaluation of the German Version of the “INTERMED-Self-Assessment”-Questionnaire (IMSA) to Assess Case Complexity]. Psychotherapie, Psychosomatik, medizinische Psychologie. 2016;66(5):180–6.

21. Cohen JS, Lyons JS, Benchimol EI, Carman N, Guertin C, Mack DR. The pediatric inflammatory bowel disease INTERMED: A new clinical tool to assess psychosocial needs. Journal of Psychosomatic Research. 2019 Apr;119:26–33.

22. Lobo E, De Jonge P, Huyse FJ, Slaets JPJ, Rabanaque M-J, Lobo A. Early Detection of Pneumology Inpatients at Risk of Extended Hospital Stay and Need for Psychosocial Treatment: Psychosomatic Medicine. 2007 Jan;69(1):99–105.

23. de Jonge P, Bauer I, Huyse FJ, Latour CHM. Medical Inpatients at Risk of Extended Hospital Stay and Poor Discharge Health Status: Detection With COMPRI and INTERMED. Psychosomatic Medicine. 2003 Jul;65(4):534–41.

24. Lobo E, de Jonge P, Huyse FJ, Rabanaque M-J, Suárez J, Lobo A. Predicción temprana de necesidades de intervención psicosocial especializada en pacientes con enfermedades pulmonares a partir de evaluaciones por enfermeras. Medicina Clínica. 2008 Nov;131(19):731–6.

25. Matzer F, Wisiak UV, Graninger M, Söllner W, Stilling HP, Glawischnig-Goschnik M, et al. Biopsychosocial Health Care Needs at the Emergency Room: Challenge of Complexity. Laks J, editor. PLoS ONE. 2012 Aug 28;7(8):e41775.

26. Koch N, Stiefel F, Jonge P de, Fransen J, Chamot A-M, Gerster J-C, et al. Identification of case complexity and increased health care utilization in patients with rheumatoid arthritis. Arthritis & Rheumatism. 2001 Jun;45(3):216–21.

27. Di Gangi Herms AMR, Pinggera GM, de Jonge P, Strasser H, Söllner W. Assessing Health Care Needs and Clinical Outcome With Urological Case Complexity: A Study Using INTERMED. Psychosomatics. 2003 May;44(3):196–203.

28. Peters LL, Burgerhof JGM, Boter H, Wild B, Buskens E, Slaets JPJ. Predictive validity of a frailty measure (GFI) and a case complexity measure (IM-E-SA) on healthcare costs in an elderly population. Journal of Psychosomatic Research. 2015 Nov;79(5):404–11.

29. de Jonge P, Ruinemans GM-F, Huyse FJ, ter Wee PM. A simple risk score predicts poor quality of life and non-survival at 1 year follow-up in dialysis patients. Nephrology Dialysis Transplantation. 2003 Dec 1;18(12):2622–8.

30. Wild B, Heider D, Maatouk I, Slaets J, König H-H, Niehoff D, et al. Significance and Costs of Complex Biopsychosocial Health Care Needs in Elderly People: Results of a Population-Based Study. Psychosomatic Medicine. 2014 Sep;76(7):497–502.

31. Eissens van der Laan MR, van Offenbeek MAG, Broekhuis H, Slaets JPJ. A personcentred segmentation study in elderly care: Towards efficient demand-driven care. Social Science & Medicine. 2014 Jul;113:68–76.

32. Wild B, Heider D, Schellberg D, Böhlen F, Schöttker B, Muhlack DC, et al. Caring for the elderly: A person-centered segmentation approach for exploring the association between health care needs, mental health care use, and costs in Germany. Hashimoto K, editor. PLOS ONE. 2019 Dec 19;14(12):e0226510.

33. de Jonge P, Latour CHM, Huyse FJ. Implementing Psychiatric Interventions on a Medical Ward: Effects on Patients’ Quality of Life and Length of Hospital Stay: Psychosomatic Medicine. 2003 Nov;65(6):997–1002.

34. Wild B, Herzog W, Schellberg D, Böhlen F, Brenner H, Saum K, et al. A short intervention targeting psychosomatic care in older adults with complex health care needs–results of a randomized controlled trial. International Journal of Geriatric Psychiatry. 2019 Feb;34(2):272–9.

35. Spoorenberg SLW, Wynia K, Uittenbroek RJ, Kremer HPH, Reijneveld SA. Effects of a population-based, person-centred and integrated care service on health, wellbeing and self-management of community-living older adults: A randomised controlled trial on Embrace. Evans CJ, editor. PLOS ONE. 2018 Jan 19;13(1):e0190751.

36. Papeix C, Gambotti L, Assouad R, Ewenczyck C, Tanguy M-L, Pineau F, et al. Evaluation of an integrated multidisciplinary approach in multiple sclerosis care: A prospective, randomized, controlled study. Multiple Sclerosis Journal - Experimental, Translational and Clinical. 2015 Jun 5;1:205521731560886.

37. Stiefel F, Zdrojewski C, Bel Hadj F, Boffa D, Dorogi Y, So A, et al. Effects of a Multifaceted Psychiatric Intervention Targeted for the Complex Medically Ill: A Randomized Controlled Trial. Psychotherapy and Psychosomatics. 2008;77(4):247–56.

38. Kathol RG, Andrew RL, Squire M, Dehnel P. The Integrated Case Management Manual: Value-Based Assistance to Complex Medical and Behavioral Health Patients [Internet]. 2nd ed. Springer International Publishing; 2018 [cited 2019 May 20]. Available from: https://www.springer.com/la/book/9783319747415

39. Barcones MF, Amatriain E, Lobo E, de la Cámara C, Lobo A. “Complex patients” in Primary Care. Journal of Psychosomatic Research. 2019 Jun;121:140.

40. Brazil. National Politics of Primary Health Care [Internet]. Ministry of Health. Department of Primary Care; 2012. Available from: http://189.28.128.100/dab/docs/publicacoes/geral/pnab.pdf.

41. Weber B. [Translation, cross-cultural adaptation and validation of the INTERMED method to the Portuguese language: study involving inpatients]. [Internet] [PhD Thesis]. [São Paulo]: University of São Paulo; 2012. Available from: https://teses.usp.br/teses/disponiveis/7/7140/tde-08102012-161016/publico/Bernardete_Weber_Tese_2012_0927.pdf

42. Weber B, Fratezi FR, Aranha Suzumura É, Ozello Gutierrez BA, De Negri Filho A, Trench Ciampone MH. [Translation and cultural adaptation of the Interdisciplinary Medicine Instrument (INTERMED): method of biopsychosocial assessment in Brazil]. RAHIS. 2012 Nov 29;(9):87.

43. Melller W, Specher S, Schultz P, Kishi Y, Thurber S, Kathol R. Using the INTERMED complexity instrument for a retrospective analysis of patients presenting with medical Illness, substance use disorder, and other psychiatric Illnesses. ANNALS OF CLINICAL PSYCHIATRY. 2015;1(27):39–43.

44. Botega NJ, Bio MR, Zomignani MA, Garcia Jr C, Pereira WAB. Mood disorders among medical in-patients: a validation study of the hospital anxiety and depression scale (HAD). Revista de Saúde Pública. 1995 Oct;29(5):359–63.

45. Griep RH, Chor D, Faerstein E, Werneck GL, Lopes CS. Construct validity of the Medical Outcomes Study’s social support scale adapted to Portuguese in the Pró-Saúde Study. Cadernos de Saúde Pública. 2005 Jun;21(3):703–14.

46. Souza RC de, Pinheiro RS, Coeli CM, Camargo Jr. KR de. The Charlson comorbidity index (CCI) for adjustment of hip fracture mortality in the elderly: analysis of the importance of recording secondary diagnoses. Cadernos de Saúde Pública. 2008 Feb;24(2):315–22.

47. World Health Organization. Division of Mental Health. WHOQOL-BREF?: introduction, administration, scoring and generic version of the assessment: field trial version. World Health Organization; 1996.

48. Lebrão ML, editor. Health, Well-Being and aging: the SABE Study in São Paulo, Brazil. 2005;8(2):127–41.

49. Salvador-Carulla L, Gonzalez-Caballero J. Assessment instruments in mental health: description and metric properties. In: Mental Health Outcome Measures. 3rd ed. RCPsych Publications; 2010. p. 28–61.

50. Harris PA, Taylor R, Minor BL, Elliott V, Fernandez M, O’Neal L, et al. The REDCap consortium: Building an international community of software platform partners. Journal of Biomedical Informatics. 2019 Jul;95:103208.

51. Dancey CP, Reidy J. Statistics without maths for psychology. 5th ed. Harlow, England?; New York: Prentice Hall/Pearson; 2011. 620 p.

52. George D, Mallery P. SPSS for Windows step by step: a simple guide and reference, 11.0 update. Boston: A & B; 2003.

53. Kishi Y, Matsuki M, Mizushima H, Matsuki H, Ohmura Y, Horikawa N. The INTERMED Japanese version: Inter-rater reliability and internal consistency. Journal of Psychosomatic Research. 2010 Dec;69(6):583–6.

54. Rudin R, Gidengil C, Predmore Z, Schneider E, Sorace J, Hornstein R. Identifying and Coordinating Care for Complex Patients: Findings from the Leading Edge of Analytics and Health Information Technology [Internet]. RAND Corporation; 2016 [cited 2020 Jun 27]. Available from: <https://www.rand.org/pubs/research_reports/RR1234.html>

55. Glasgow RE, Kaplan RM, Ockene JK, Fisher EB, Emmons KM. Patient-Reported Measures Of Psychosocial Issues And Health Behavior Should Be Added To Electronic Health Records. Health Affairs. 2012 Mar;31(3):497–504.

56. Senteio C, Adler-Milstein J, Richardson C, Veinot T. Psychosocial information use for clinical decisions in diabetes care. Journal of the American Medical Informatics Association. 2019 Aug 1;26(8–9):813–24.

57. Munchhof A, Gruber R, Lane KA, Bo N, Rattray NA. Beyond Discharge Summaries: Communication Preferences in Care Transitions Between Hospitalists and Primary Care Providers Using Electronic Medical Records. Journal of General Internal Medicine. 2020 Jun;35(6):1789–96.

58. Imprinting Care and the Loss of Patient Narrative: Creation and Standardization of Medical Records. The Permanente Journal [Internet]. 2019 [cited 2020 Sep 7]; Available from: http://www.thepermanentejournal.org/issues/2019/summer/7187-psychiatric-asylum-medical-records.html

59. with the HITEC Investigators, Kern LM, Barrón Y, Dhopeshwarkar RV, Edwards A, Kaushal R. Electronic Health Records and Ambulatory Quality of Care. Journal of General Internal Medicine. 2013 Apr;28(4):496–503.

60. Brasil M da SaúdeS de P de S. Portaria n. 1101, de 12 de junho de 2002. G.1) Capacidade de produção, em consultas, de alguns recursos humanos na área de saúde [Internet]. Ministério da Saúde; 2002 [cited 2020 Jun 13]. Available from: http://www.saude.gov.br/images/pdf/2015/outubro/02/Portaria1101-2002.pdf

61. Irving G, Neves AL, Dambha-Miller H, Oishi A, Tagashira H, Verho A, et al. International variations in primary care physician consultation time: a systematic review of 67 countries. BMJ Open. 2017 Oct;7(10):e017902.

62. Berntsen G, Høyem A, Lettrem I, Ruland C, Rumpsfeld M, Gammon D. A person-centered integrated care quality framework, based on a qualitative study of patients’ evaluation of care in light of chronic care ideals. BMC Health Services Research [Internet]. 2018 Dec [cited 2020 Jun 15];18(1). Available from: https://bmchealthservres.biomedcentral.com/articles/10.1186/s12913-018-3246-z

63. Edwards ST, Dorr DA, Landon BE. Can Personalized Care Planning Improve Primary Care? JAMA. 2017 Jul 4;318(1):25.

64. Joint Action CHRODIS. Report on care pathways approaches for multimorbid chronic patients - Addressing chronic disease and healthy ageing across the life cycle. CHRODIS; 2017.

65. NICE guideline. Multimorbidity: clinical assessment and management. UK: National Institute for Health and Care Excellence, National Guideline Centre; 2016.

66. Simmons LA, Drake CD, Gaudet TW, Snyderman R. Personalized Health Planning in Primary Care Settings. Fed Pract. 2016 Jan;33(1):27–34.

67. Grant RW, Ashburner JM, Hong CS, Chang Y, Barry MJ, Atlas SJ. Defining patient complexity from the primary care physician’s perspective: A Cohort Study. Annals of Internal Medicine. 2011 Dec 20;155(12):797.

68. Muth C, van den Akker M, Blom JW, Mallen CD, Rochon J, Schellevis FG, et al. The Ariadne principles: how to handle multimorbidity in primary care consultations. BMC Medicine [Internet]. 2014 Dec [cited 2020 Jun 15];12(1). Available from: http://bmcmedicine.biomedcentral.com/articles/10.1186/s12916-014-0223-1

69. Webster F, Rice K, Bhattacharyya O, Katz J, Oosenbrug E, Upshur R. The mismeasurement of complexity: provider narratives of patients with complex needs in primary care settings. International Journal for Equity in Health [Internet]. 2019 Dec [cited 2020 Jun 27];18(1). Available from: https://equityhealthj.biomedcentral.com/articles/10.1186/s12939-019-1010-6

70. Maciejewski ML, Powers BJ, Sanders LL, Farley JF, Hansen RA, Sleath B, et al. The Intersection of Patient Complexity, Prescriber Continuity and Acute Care Utilization. Journal of General Internal Medicine. 2014 Apr;29(4):594–601.

71. Bolton RE, Bokhour BG, Hogan TP, Luger TM, Ruben M, Fix GM. Integrating Personalized Care Planning into Primary Care: a Multiple-Case Study of Early Adopting Patient-Centered Medical Homes. Journal of General Internal Medicine. 2020 Feb;35(2):428–36.

72. Berntsen GKR, Dalbakk M, Hurley JS, Bergmo T, Solbakken B, Spansvoll L, et al. Person-centred, integrated and pro-active care for multi-morbid elderly with advanced care needs: a propensity score-matched controlled trial. BMC Health Services Research [Internet]. 2019 Dec [cited 2020 Jun 15];19(1). Available from: https://bmchealthservres.biomedcentral.com/articles/10.1186/s12913-019-4397-2

73. Olmos-Ochoa TT, Bharath P, Ganz DA, Noël PH, Chawla N, Barnard JM, et al. Staff Perspectives on Primary Care Teams as De Facto “Hubs” for Care Coordination in VA: a Qualitative Study. Journal of General Internal Medicine. 2019 May;34(S1):82–9.

74. Krist AH, O’Loughlin K, Woolf SH, Sabo RT, Hinesley J, Kuzel AJ, et al. Enhanced care planning and clinical-community linkages versus usual care to address basic needs of patients with multiple chronic conditions: a clinician-level randomized controlled trial. Trials [Internet]. 2020 Dec [cited 2020 Jun 15];21(1). Available from: https://trialsjournal.biomedcentral.com/articles/10.1186/s13063-020-04463-3

75. Michelsen K, Brand H, Achterberg PW, Wilkinson JR, Health Evidence Network, World Health Organization, et al. Promoting better integration of health information systems: best practices and challenges. 2015.

76. Reed ME, Huang J, Parikh R, Millman A, Ballard DW, Barr I, et al. Patient–Provider Video Telemedicine Integrated With Clinical Care: Patient Experiences. Annals of Internal Medicine. 2019 Aug 6;171(3):222.

77. Hilty DM, Rabinowitz T, McCarron RM, Katzelnick DJ, Chang T, Bauer AM, et al. An Update on Telepsychiatry and How It Can Leverage Collaborative, Stepped, and Integrated Services to Primary Care. Psychosomatics. 2018 May;59(3):227–50.

78. World Health Organization. WHO guideline: recommendations on digital interventions for health system strengthening. World Health Organization; 2019.

79. The Topol Review. Preparing the healthcare workforce to deliver the digital future. An independent report on behalf of the Secretary of State for Health and Social Care [Internet]. NHS Health Education England; 2019. Available from: https://topol.hee.nhs.uk/

80. Borrell-Carrio F. The biopsychosocial model 25 years later: principles, practice, and scientific inquiry. The Annals of Family Medicine. 2004 Nov 1;2(6):576–82.

81. Epstein RM, Fiscella K, Lesser CS, Stange KC. Why The Nation Needs A Policy Push On Patient-Centered Health Care. Health Affairs. 2010 Aug;29(8):1489–95.

82. Heyeres M, McCalman J, Tsey K, Kinchin I. The Complexity of Health Service Integration: A Review of Reviews. Frontiers in Public Health [Internet]. 2016 Oct 17 [cited 2020 May 14];4. Available from: http://journal.frontiersin.org/article/10.3389/fpubh.2016.00223/full

83. Mounier-Jack S, Mayhew SH, Mays N. Integrated care: learning between high-income, and low- and middle-income country health systems. Health Policy and Planning. 2017 Nov 1;32(Suppl_4):iv6–12.

84. Goodyear-Smith F, Bazemore A, Coffman M, Fortier R, Howe A, Kidd M, et al. Primary Care Research Priorities in Low-and Middle-Income Countries. The Annals of Family Medicine. 2019 Jan;17(1):31–5.

